# A recurrent mutation at position 26,340 of SARS-CoV-2 is associated with failure of the E-gene qRT-PCR utilised in a commercial dual-target diagnostic assay

**DOI:** 10.1101/2020.04.28.20083337

**Authors:** Maria Artesi, Sébastien Bontems, Paul Göbbels, Marc Franckh, Piet Maes, Raphaël Boreux, Cécile Meex, Pierrette Melin, Marie-Pierre Hayette, Vincent Bours, Keith Durkin

**Affiliations:** Laboratory of Human Genetics, GIGA Research Institute, Liège, Belgium; Department of Clinical Microbiology, University Hospital of Liège, Liège, Belgium; St-Nikolaus Hospital, Eupen, Belgium; Department of Microbiology, Immunology and Transplantation, Rega Institute, KU Leuven, Leuven, Belgium; Department of Human Genetics, University Hospital of Liège, Liège, Belgium

**Author notes:** These authors contributed equally (Maria Artesi, Sébastien Bontems). These authors jointly supervised the work (Marie-Pierre Hayette, Vincent Bours, Keith Durkin).

## Abstract

Control of the ongoing severe acute respiratory syndrome coronavirus 2 (SARS-CoV-2) pandemic requires accurate laboratory testing to identify infected individuals, while also clearing essential staff to continue work. At the current time a number of qRT-PCR assays have been developed to identify SARS-CoV-2, targeting multiple positions in the viral genome. While the mutation rate of SARS-CoV-2 is moderate, given the large number of transmission chains it is prudent to monitor circulating viruses for variants that might compromise these assays. Here we report the identification of a C-to-T transition at position 26,340 of the SARS-CoV-2 genome which is associated with failure of the cobas® SARS-CoV-2 E-gene qRT-PCR in eight patients. As the cobas® SARS-CoV-2 assay targets two positions in the genome, the individuals carrying this variant were still called as SARS-CoV-2 positive. Whole genome sequencing of SARS-CoV-2 showed all to carry closely related viruses. Examination of viral genomes deposited on GISAID showed this mutation has arisen independently at least four times. This work highlights the necessity of monitoring SARS-CoV-2 for the emergence of SNPs which might adversely affect RT-PCRs used in diagnostics. Additionally, it argues that two regions in SARS-CoV-2 should be targeted to avoid false negatives.

## Introduction

Coronavirus disease of 2019 (COVID-19) originated in Wuhan, China in late 2019, [1,2] generating a global pandemic [3]. As of the 22th of May 2020, there have been close to 5 million confirmed cases and more than 300,000 deaths reported worldwide [4]. Metagenomic RNA sequencing revealed that COVID-19 is caused by a novel coronavirus, subsequently named severe acute respiratory syndrome coronavirus (SARS-CoV-2). SARS-CoV-2 is a close relative of SARS-CoV and MERS-CoV [2], coronaviruses which have both been responsible for large outbreaks of respiratory illness within the last two decades [5,6]. The release of the first SARS-CoV-2 genome sequence on the 10th of January spurred the development of RT-PCR assays [7–9] and thereby enabled reliable laboratory diagnosis of infections. In addition, protocols have been developed to allow for rapid sequencing of the SARS-CoV-2 genome [10], sharing of the resultant data [11] and phylogenetic analysis [12,13].

Laboratory testing for SARS-CoV-2 is a cornerstone of the strategy to mitigate its spread as it facilitates the identification and isolation of infected individuals, while negative tests can allow essential personnel to continue work [14]. In the context of SARS-CoV-2, due to its high transmissibility [15], false negatives could have particularly adverse effects on efforts to control its spread. As qRT-PCR oligos rely on binding to small ∼20bp regions, mutations in these targets have the potential to impair efficient amplification or probe binding, thereby generating false negatives. In contrast to other RNA viruses, coronaviruses have a moderate mutation rate due their ability to carry out RNA proofreading [16]. Nevertheless, given the large number of ongoing transmission chains, it remains prudent to monitor the integrity of qRT-PCR assays.

Here we report the identification of a SNP in the E-gene of SARS-CoV-2 that is associated with the failure of the qRT-PCR which targets the E-gene in the cobas® SARS-CoV-2 test (Roche). As this dual-target assay also detects a region in ORF1b these samples were still correctly identified as SARS-CoV-2 positive. This observation highlights the necessity of targeting two regions in SARS-CoV-2 RT-PCR assays and shows the role sequencing can play in resolving and anticipating problems with the qRT-PCR assays in use.

## Methods

### RNA extraction and Real Time PCR

The study was approved by the Comité d’Ethique Hospitalo-Facultaire Universitaire de Liège (Reference number: CE 2020/137). The COVID-19 detection was routinely performed using the cobas® 6800 platform (Roche). For this, 400 µL of nasopharyngeal swabs in a preservative medium (AMIES or UTM) were first incubated at room temperature for 30 minutes with 400 µL of cobas ® PCR Media kit (Roche) for viral inactivation. Samples were then loaded on the cobas® 6800 platform using the cobas® SARS-CoV-2 assay for the detection of the ORF1ab and E genes.

For qRT-PCR control and sequencing analysis, RNA was extracted from clinical samples (300µL) on a Maxwell 48 device using the Maxwell RSC Viral RNA kit (Promega) following a viral inactivation step using Proteinase K according to manufacturer’s instructions. RNA elution occurred in 50µL RNAse free water and 5 µL were used for the RT-PCR. Reverse transcription and RT-PCR were performed on a LC480 thermocycler (Roche) based on Corman et al. [9] protocol for the detection of RdRP and E genes using the Taqman Fast Virus 1-Step Master Mix (Thermo Fisher). Primers and probes (Eurogentec, Belgium) were used as described by the authors [9].

### SARS-CoV-2 whole genome sequencing

Reverse transcription was carried out using SuperScript IV VILO™ Master Mix, 3.3 µL of RNA was combined with 1.2 µL of master mix and 1.5 µL of H_2_O. This was incubated at 25°C for 10 min, 50°C for 10 min and 85°C for 5 min. PCR reactions used the primers and conditions recommended in the nCoV-2019 sequencing protocol [17]. Version 3 of the Artic network primers were used, these were synthesised by Integrated DNA Technologies. Samples were multiplexed using the Oxford Nanopore Native Barcoding Expansion kits 1-12 and 13-24, in conjunction with Ligation Sequencing Kit. Sequencing was carried out on a Minion using R9.4.1 flow cells. Data analysis followed the nCoV-2019 novel coronavirus bioinformatics protocol of the Artic network [17]. The resulting consensus viral genomes have been deposited at the Global Initiative on Sharing All Influenza Data (GISAID)[11]

### Sanger sequencing

Reverse transcription was carried out as above. The primers nCoV-2019_87_LEFT and nCoV-2019_87_RIGHT from the Artic network nCoV-2019 amplicon set [17] were used to amplify the regions between positions 26198-26590. The resultant PCR product was purified using Ampure XP beads (Beckman Coulter), sequenced using BigDye terminator cycle-sequencing kit (Applied Biosystems) and run on a ABI PRISM 3730 DNA analyser (Applied Biosystems).

### Phylogeny

SARS-CoV-2 genomes and the associated metadata were downloaded from GISAID (https://www.gisaid.org/) on the 25th May 2020. Viral genomes marked by GISAID as complete (>29,000 bases) and high coverage (<1% Ns, <0.05% unique amino acid mutations and no insertion/deletions unless verified by submitter) were selected, leaving 20,386 viral genomes. We also download viral genomes using a less stringent cut off, requiring the virus to be complete (>29,000) and excluding viruses with low coverage (>5% Ns), in this case 29,699 viral genomes remained. Viruses carrying a variant at position 26,340 were identified with SeqKit [18] using the following grep command and motif encompassing the variant (underlined) “seqkit grep -s -i -p TTACACTAGC<u>T</u>ATCCTTACTG”. The viruses containing the variant were added to the list of viruses to include in the Nextstrain build. Viruses from non-human hosts were excluded from the analysis. Nextstrain phylogenetic trees were generated for both data sets using the default configuration (https://github.com/nextstrain/ncov). The SARS-CoV-2 genomes were assigned to a lineage via pangolin (https://github.com/hCoV-2019/pangolin) which used the virus nomenclature proposed by Rambaut et al. 2020 [19].

## Results

The cobas® system (Roche) implements a dual target assay to detect SARS-CoV-2, with qRT-PCRs targeting both the ORF1ab region and the E-gene (Supplementary Figure 1). During the course of routine SARS-CoV-2 testing, we observed eight samples that were negative for the E-gene qRT-PCR, but positive for the ORF1ab qRT-PCR (Table 1). Four of these samples were retested using the Corman et al. [9] SARS-CoV-2 assay that targets the RdRP and E genes. In this instance both qRT-PCRs were positive (Table 1). All came from Belgian healthcare workers in the same service, with sampling dates that ranged between 23rd March and 17th April 2020 (Figure 1 A). As the samples were positive for the ORF1ab qRT-PCR all samples were correctly classified as positive by the cobas® system (Roche).

**Table 1.** Real-time PCR Ct values observed in the 8 patients, two patients were tested twice, two weeks apart (+Initial test, *2-week test). Four of the samples (underlined) were also tested using the SARS-CoV-2 assay of Corman et al. 2020.

|  | cobas® |  | Corman et al. 2020 |  |
| --- | --- | --- | --- | --- |
| patient ID | RdRp (Ct) | E gene (Ct) | Ct RdRp | Ct E gene |
| ULG-0184 | 21.53 | 0 | nd | nd |
| <u>ULG-10088</u> | <u>26.65</u> | <u>0</u> | <u>28.66</u> | <u>26.51</u> |
| <u>ULG-10089</u> | <u>22.64</u> | <u>0</u> | <u>25.02</u> | <u>22.85</u> |
| <u>ULG-10095+</u> | <u>24.62</u> | <u>0</u> | <u>23.73</u> | <u>22</u> |
| ULG-10095* | 31.94 | 0 | nd | nd |
| <u>ULG-10096</u> | <u>20.45</u> | <u>0</u> | <u>22.89</u> | <u>20.33</u> |
| ULG-10169+ | 28.64 | 0 | nd | nd |
| ULG-10169* | 28.55 | 0 | nd | nd |
| ULG-10145 | 27.19 | 0 | nd | nd |
| ULG-10146 | 25.44 | 0 | nd | nd |

**Figure 1.**
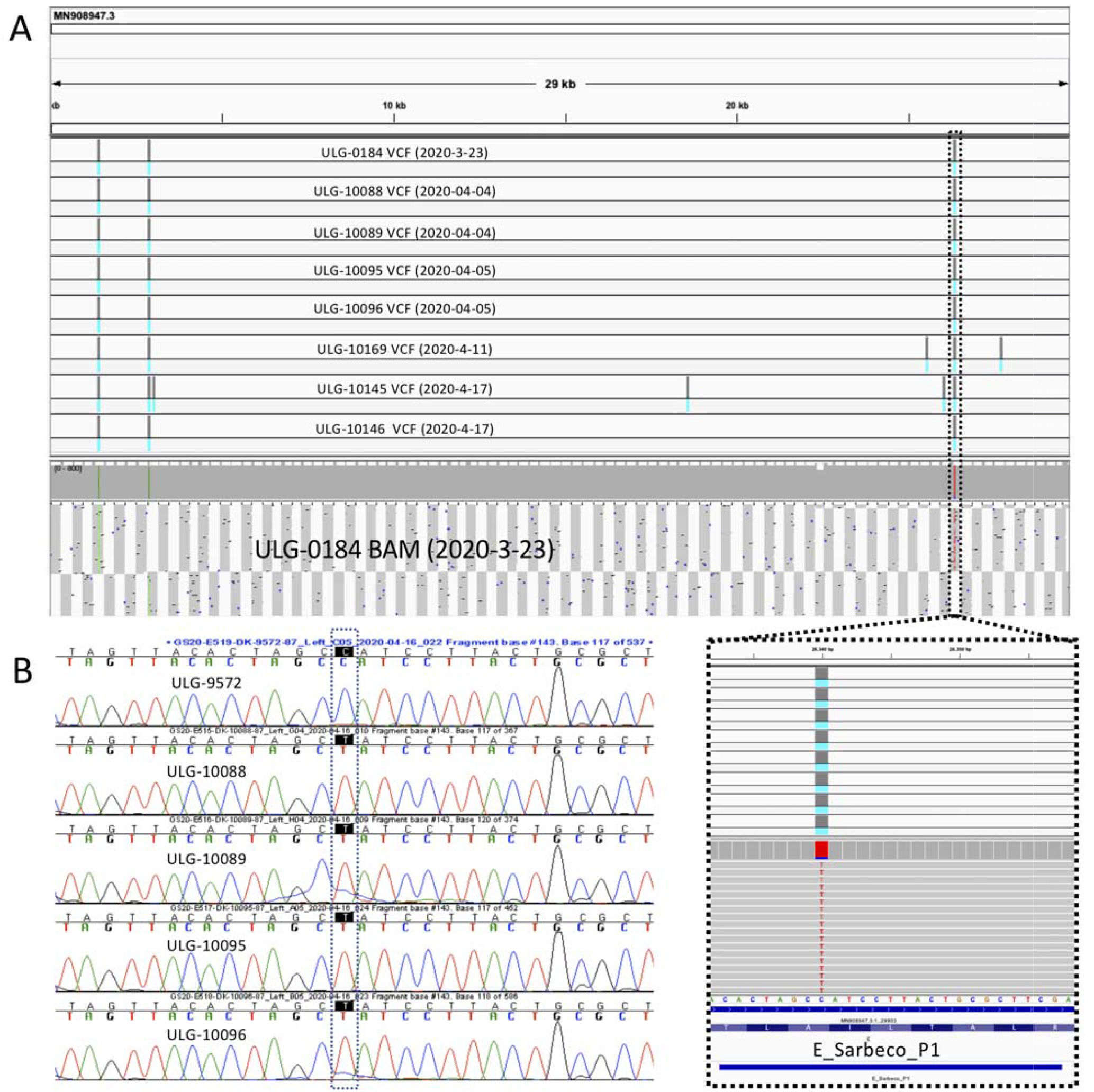
**(A)** Screen shot form IGV (Integrative Genomics Viewer: http://software.broadinstitute.org/software/igv/home) shows the VCFs (lines represent SNPs) for the eight viruses as well as a BAM files showing the reads and coverage for one virus. Sampling date for each virus is indicated. The zoomed section shows the SNP at 26,340, the blue rectangle labelled E_Sarbeco_P1 corresponds to the region covered by the E gene probe in Corman et al. [9]. **(B)** Sanger sequencing of four samples carrying the C-to-T transition at position 26,340. The top chromatogram shows a virus carrying the wildtype sequence.

We speculated that these samples carried a common variant that interfered with the E-gene qRT-PCR and carried out whole genome sequencing of the viruses using the Artic Network protocol [17]. The consensus genomes generated showed six to be infected with a genetically identical virus (Figure 1A). The remaining two viruses shared the same SNPs as the previous six, but had accumulated additional mutations (suggesting continued spread of the lineage in the area). In two cases we also had a two week follow up sample from the same patient, in each case the consensus viral genomes generated were identical (Supplementary Figure 2). The six identical viruses (derived from different patients) deviated from the MN908947.3 reference isolated in Wuhan at only three positions (Figure 1 A). The first two SNPs were towards the 5’ end of the virus at positions 1,440 and 2,891 respectively. The third SNP, a C-to-T transition at position 26,340 is within the E gene of the virus and was validated by Sanger sequencing in four samples (Figure 1 B). This SNP overlaps with the E gene probe used in the Corman et al. [9] RT-PCR assay, however, as was mentioned above, it does not appear to affect the performance of this assay in our hands. Unfortunately, the position of primers and probes utilised in the cobas® E-gene assay (Roche) are not publicly available, nevertheless it is parsimonious to assume that this SNP is the cause of the failure of the E-gene qRT-PCR implemented in the cobas® system.

Out of the 229 SARS-CoV-2 genomes we have sequenced at the time of writing, eight carry the SNP at position 26,340. To see if the same variant was circulating more widely we examined the SARS-CoV-2 sequences deposited in GISAID for a variant at the same position. When only complete, high coverage genomes are considered (20,386 genomes), eighteen were found to carry a C-to-T transition at position 26,340. Eight of these were sequenced by us, seven were isolated in England, two in Switzerland and one in Turkey. As can be seen in Figure 2, viruses isolated in the same country cluster together, however they do not cluster with other viruses carrying the SNP at position 26,340. We also classified the viral genomes according to the nomenclature proposed by Rambaut et al. 2020 [19]. Table 2 shows that samples isolated in the same country belong to the same lineage, with no overlap in lineage between countries. As a consequence, it appears that this variant has arisen multiple times in different transmission chains (homoplasic site).

**Figure 2.**
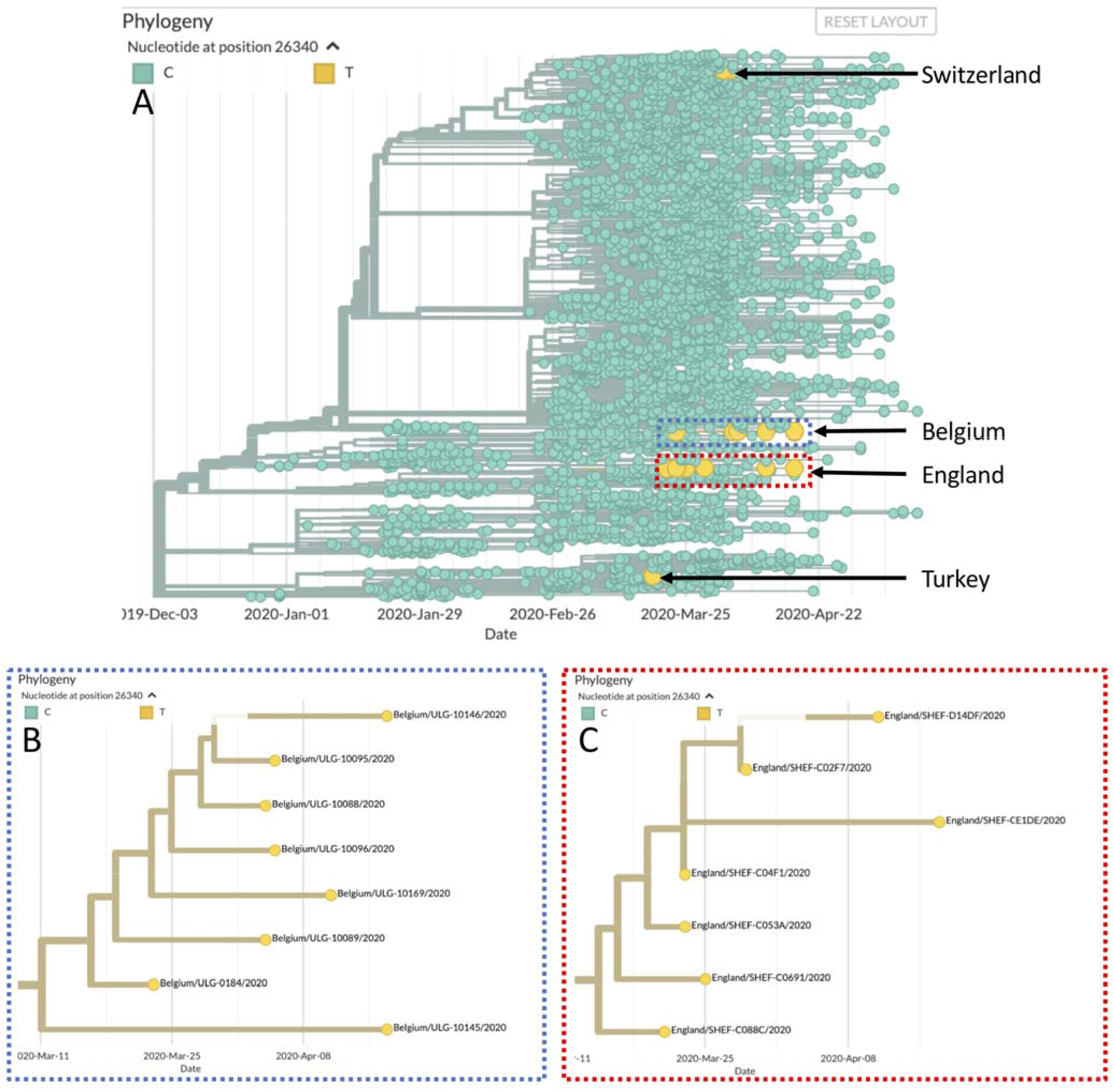
**(A)** Phylogenetic tree generated by Nextstrain [12] with the viruses carrying the T allele at 26,340 highlighted in yellow. (**B)** Expanded view of the eight Belgian samples (**C)** Expanded view of the seven English samples.

**Table 2.** Lineages of the high coverage viruses, using the scheme of Rambaut et al. 2020 [19]

| Virus name | Lineage | Virus name | Lineage |
| --- | --- | --- | --- |
| Switzerland/GE0304/2020 | B.1.5 | Belgium/ULG-0184/2020 | B.3 |
| Switzerland/GE2453/2020 | B.1.5 | Belgium/ULG-10088/2020 | B.3 |
| England/SHEF-C02F7/2020 | B.2.1 | Belgium/ULG-10089/2020 | B.3 |
| England/SHEF-C04F1/2020 | B.2.1 | Belgium/ULG-10095/2020 | B.3 |
| England/SHEF-C053A/2020 | B.2.1 | Belgium/ULG-10096/2020 | B.3 |
| England/SHEF-C0691/2020 | B.2.1 | Belgium/ULG-10145/2020 | B.3 |
| England/SHEF-C088C/2020 | B.2.1 | Belgium/ULG-10146/2020 | B.3 |
| England/SHEF-CE1DE/2020 | B.2.1 | Belgium/ULG-10169/2020 | B.3 |
| England/SHEF-D14DF/2020 | B.2.1 | Turkey/HSGM-8992/2020 | B.4 |

Finally, we relaxed the filtering of viral genomes, selecting genomes >29,000 bases in length and with less than 5% Ns (we no longer required the virus to be classified as high coverage). This added 9,313 genomes (29,699 in total) and revealed eight additional viruses carrying a C-to-T transition at 26,340 (Supplementary Figure 3). Of these six were isolated in England, four clustered with the previous English samples, while the other two fell in different parts of the tree. Of the remaining two viruses, one was isolated in Australia and the second was sequenced in Luxembourg. Interestingly the Luxembourg virus clustered with the samples identified by us and was assigned to the same B.3 lineage, suggesting it to be part of the same cluster of infections.

## Discussion

As the positions of the primers and probes used in the cobas® (Roche) E-gene qRT-PCR have not been disclosed to us upon request, we cannot definitively conclude that the C-to-T transition at position 26,340 of the SARS-CoV-2 genome causes the failure in the E-gene qRT-PCR in the patients examined. However, given the available data, causality appears likely. The cobas® E-gene qRT-PCR may use an alternate primer probe combination that is more sensitive to the presence of the SNP, alternatively it may target the same positions as the Corman et al. [9] E-gene assay, but differences in reagents used and cycling conditions may prevent binding of the probe in the presence of the SNP.

It should be stressed that despite the failure of the E-gene qRT-PCR in these patients the cobas® assay correctly called these individuals as positive for SARS-CoV-2 due to a positive ORF1ab qRT-PCR. This highlights the prudence of targeting more than one position in the viral genome in a diagnostic assay. The Corman et al. [9] protocol recommends the use of its E-gene assay as a first-line screening tool, with confirmatory testing using the RdRp gene assay [9]. This SNP does not affect the Corman et al. [9] E-gene qRT-PCR in our hands, however our results highlight how a mutation in the virus can generate a false negative in a single qRT-PCR. In most cases these mutations will be rare, however as our examination of the GISAID data has shown, such mutations have the potential to arise independently in separate transmission chains.

Recently Vogels et al [20] examined the efficiency as well as frequency of variants impacting a number of the qRT-PCRs commonly used for SARS-CoV-2 testing. They found a number of variants that fell within the primer and probe binding sites, with the majority present at a low frequency and involving only a single base. A prominent exception involved a GGG→AAC mutation at genome positions 28,881-28,883 that overlaps with the first three bases of the 5’ end of the Chinese CDC N gene forward primer [7]. This mutation is found in approximately 25% of the viruses on GISAID (accessed 25th May 2020). Given the high frequency of this variant it would appear prudent to avoid using this qRT-PCR primer.

This work shows the danger of relying on an assay targeting a single position in the viral genome. It also highlights the utility of combining testing with rapid sequencing of a subset of the positive samples, especially in cases where one the qRT-PCRs fails. The sequencing allowed us to pinpoint the likely reason behind the failure of the E gene qRT-PCR. The identification of viruses carrying additional mutations as well as the clustering of the Luxembourg virus with the Belgian viruses also suggests that only a fraction of the virus carrying this variant came to our attention. This highlights that, while the variant is at a low frequency globally, at the local level it could be much higher. This example shows that it remains prudent to continue monitoring viral genomes for variants that can negatively impact this and other diagnostic assays. Finally, it would be preferable if manufacturers were transparent about the primer and probes used, this would allow problematic variants to be more readily identified from the available viral sequences.

## Data Availability

All the viral genomes have been deposited at GISAID

## Author contributions

M.A. carried out SARS-CoV-2 sequencing, S.B., R.B. and C.M. performed RNA extraction and qRT-PCR assays. qRT-PCR assays. P.G. and M.F. collected patient samples and information. P.M. (KU Leuven) assisted with SARS-CoV-2 sequencing. P.M. (Liège) & MP.H. supervised and guided the work in the Department of Clinical Microbiology. V.B. supervised and guided the work in the Laboratory of Human Genetics. K.D. carried out SARS-CoV-2 sequencing, data analysis and wrote the manuscript, with input from the other authors.

## Acknowledgements

This work was supported by the Région Wallonne project WALGEMED (convention n° 1710180). We would like to acknowledge and thank the laboratories who submitted and shared their sequences to GISAID. We would also like to thank the members of the GIGA-Genomic platform for the Sanger sequencing. Thanks to Lize Cuypers (KU Leuven) for carrying out conformation qRT-PCR assays. Finally thanks to Josh Quick (University of Birmingham) for providing an aliquot of the V2 Artic network primers.

---

**Supplementary Figure 1.**
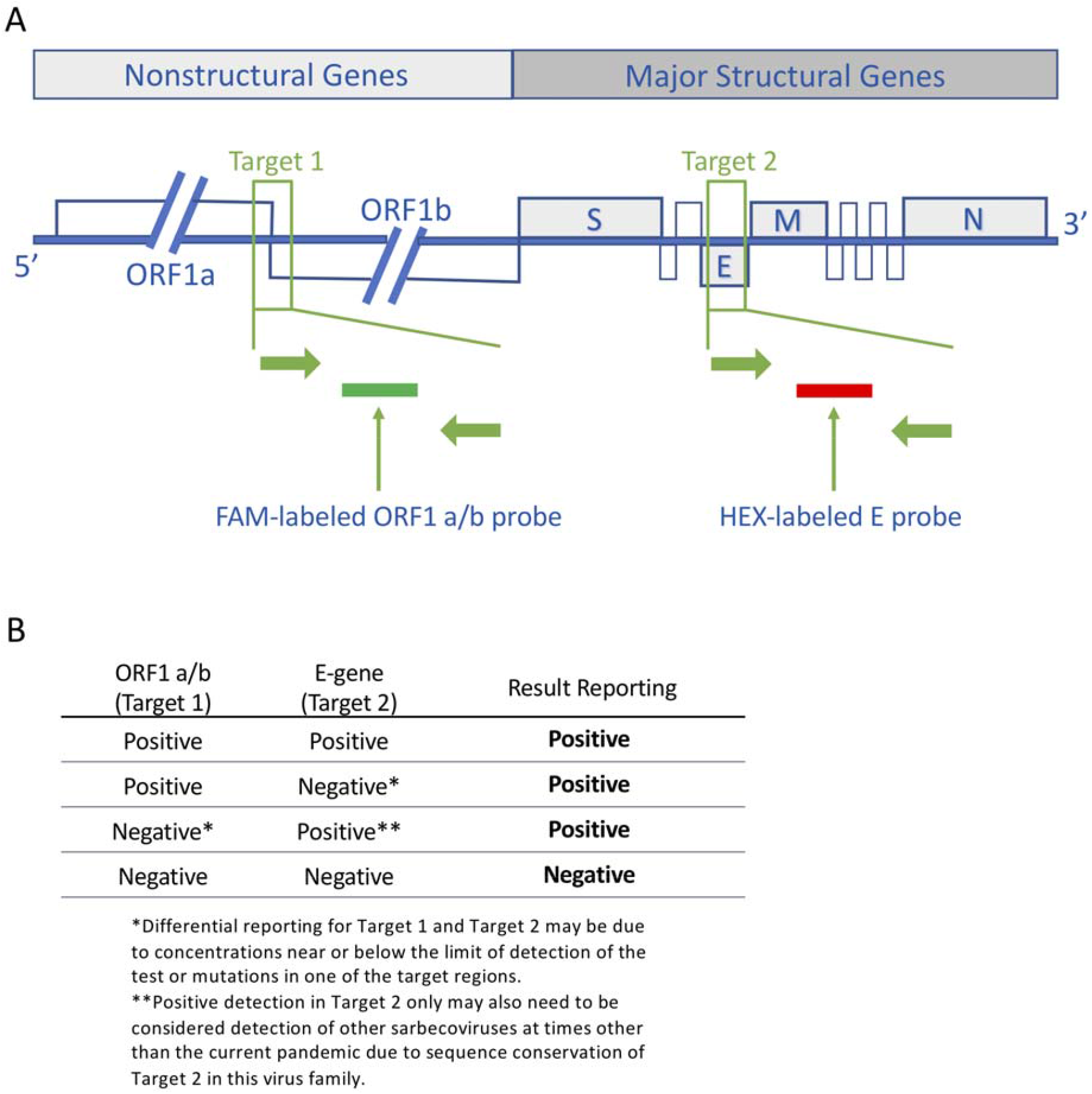
**(A)** SARS-CoV-2 genome and locations of target regions for the cobas® SARS-CoV-2 Test. The test contains a dual-target design with primers and detection probes which are specific for the ORF1 a/b non-structural region that is unique to SARS-CoV-2 and a conserved region in the structural protein envelope E-gene for SARS-CoV-1 and SARS-CoV-2 detection. Accurate detection of novel SARS-CoV-2 can be impaired by primer and probe target sequence polymorphisms and a dual-target amplification approach was realized to cope with the genetic diversity and ongoing evolution of the virus. **(B)** Test interpretation based on target results ORF: open reading frame, E: envelope protein gene, M: membrane protein gene, N: nucleocapsid protein gene, S: spike protein gene (This figure was provided by Roche)

**Supplementary Figure 2.**
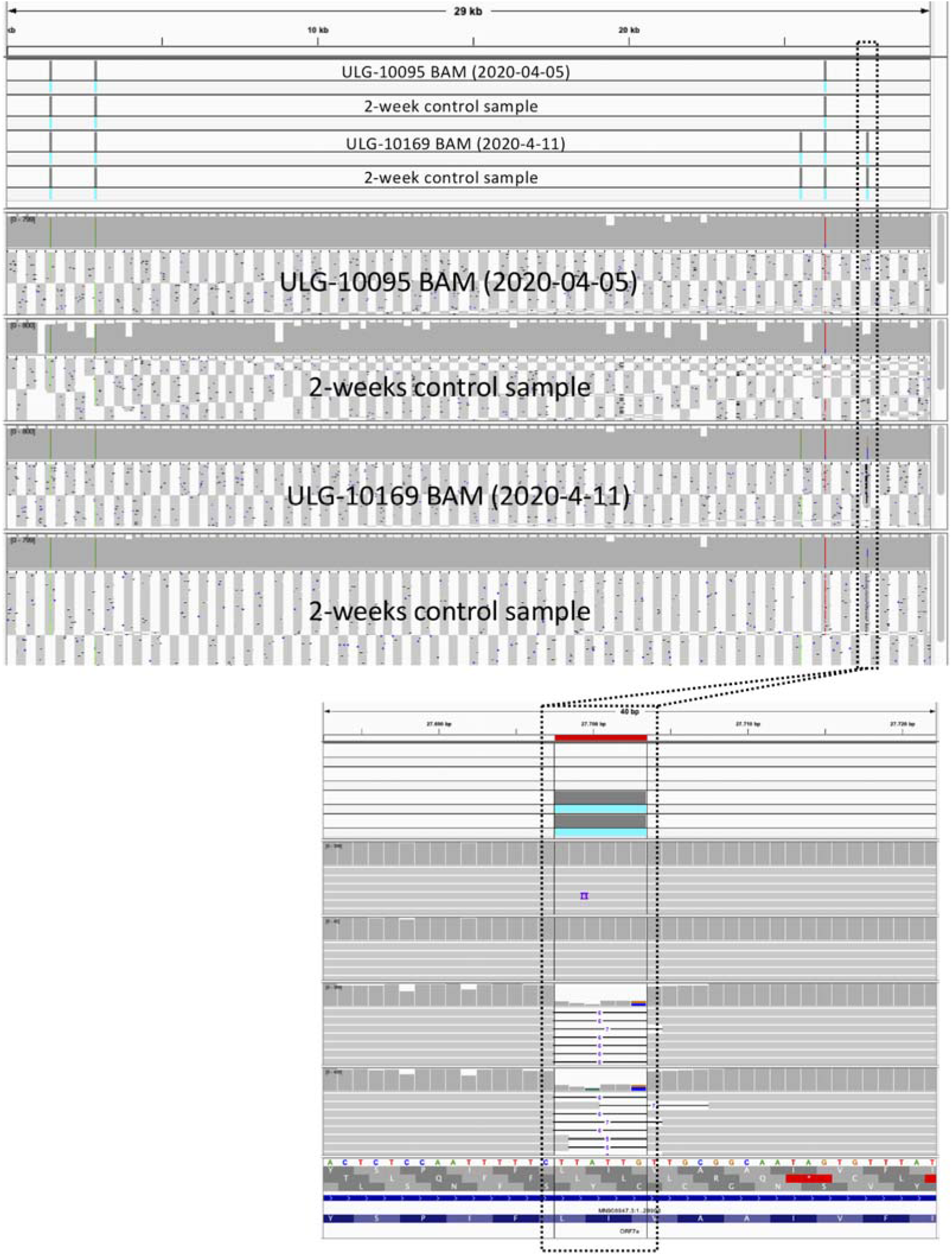
Screen shot form IGV shows VCFs and BAMs from the two patients that were sampled two weeks apart. Samples from both time points were identical. The six-base deletion (Position: 27,697-27,703) in patient ULG-10169 was observed in both time points.

**Supplementary Figure 3.**
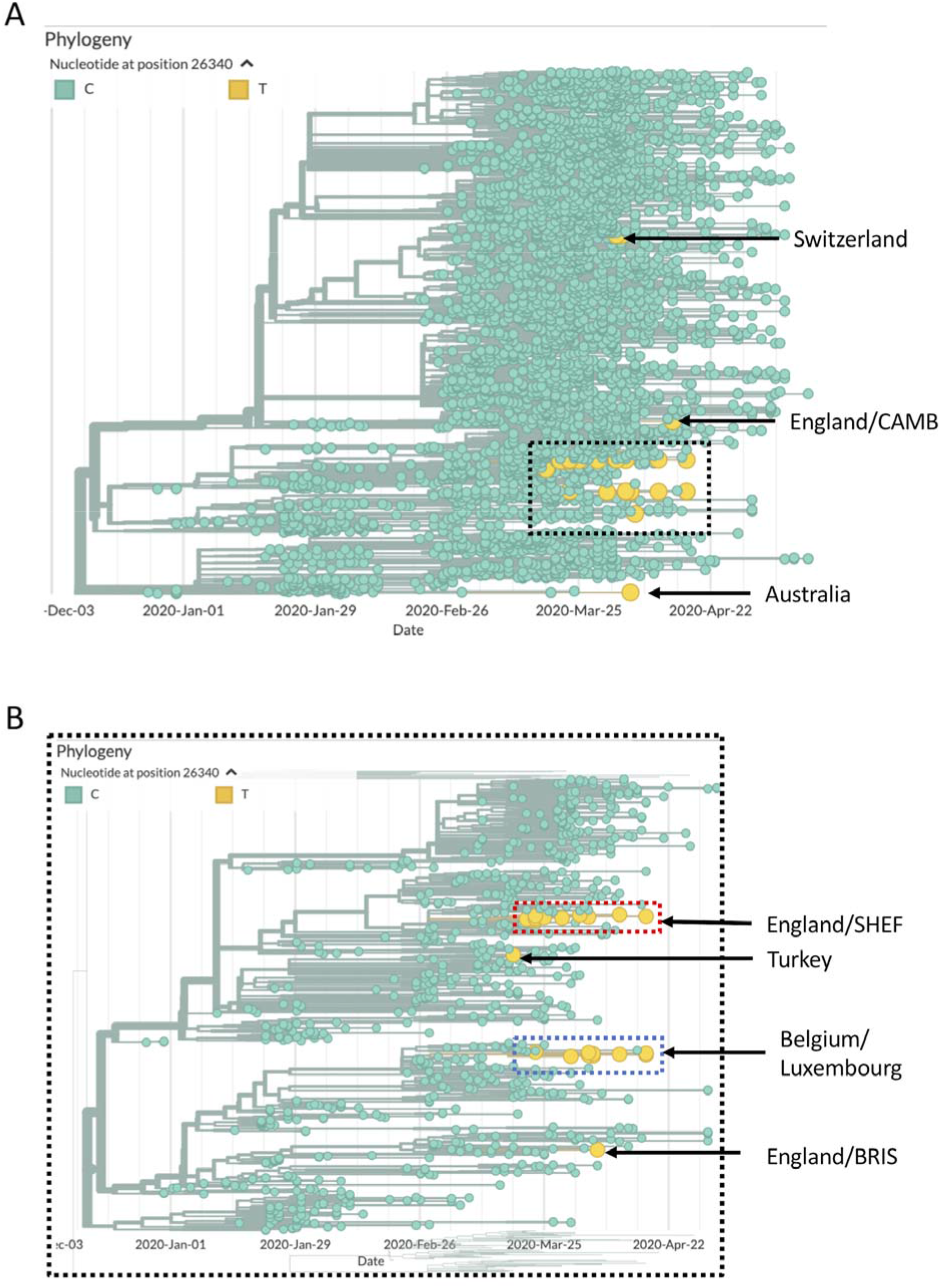
**(A)** Phylogenetic tree generated by Nextstrain [12] with the 26 viruses (low coverage excluded) carrying the T allele at 26,340 highlighted in yellow. (**B)** Expanded view of the part of the tree with the Belgian/Luxembourg, English SNHEF/BRIS and Turkish samples.

**Supplementary Table 1.** The sequence from the 26 SARS-CoV-2 genomes (low coverage excluded) carrying the T allele at 26,340 were analysed with the Pangolin software package. It assigns SARS-CoV-2 genome sequences to global lineages proposed by Rambaut et al. 2020.

| Virus name | lineage | SH-alrt | UFbootstra<br>p | lineages_version | status |
| --- | --- | --- | --- | --- | --- |
| England/BRIS-123640/2020 | B | 100 | 100 | 07/05/2020 | passed_qc |
| England/CAMB-82E84/2020 | B.1 | 100 | 100 | 07/05/2020 | passed_qc |
| Switzerland/GE0304/2020* | B.1.5 | 100 | 97 | 07/05/2020 | passed_qc |
| Switzerland/GE2453/2020* | B.1.5 | 100 | 100 | 07/05/2020 | passed_qc |
| England/SHEF-C02F7/2020* | B.2.1 | 100 | 100 | 07/05/2020 | passed_qc |
| England/SHEF-C04F1/2020* | B.2.1 | 100 | 98 | 07/05/2020 | passed_qc |
| England/SHEF-C053A/2020* | B.2.1 | 100 | 98 | 07/05/2020 | passed_qc |
| England/SHEF-C0691/2020* | B.2.1 | 100 | 97 | 07/05/2020 | passed_qc |
| England/SHEF-C088C/2020* | B.2.1 | 100 | 99 | 07/05/2020 | passed_qc |
| England/SHEF-C2B45/2020 | B.2.1 | 100 | 96 | 07/05/2020 | passed_qc |
| England/SHEF-C3090/2020 | B.2.1 | 100 | 97 | 07/05/2020 | passed_qc |
| England/SHEF-C9883/2020 | B.2.1 | 100 | 97 | 07/05/2020 | passed_qc |
| England/SHEF-CE1DE/2020* | B.2.1 | 100 | 97 | 07/05/2020 | passed_qc |
| England/SHEF-CE684/2020 | B.2.1 | 100 | 98 | 07/05/2020 | passed_qc |
| England/SHEF-D14DF/2020* | B.2.1 | 100 | 97 | 07/05/2020 | passed_qc |
| Belgium/ULG-0184/2020* | B.3 | 100 | 95 | 07/05/2020 | passed_qc |
| Belgium/ULG-10088/2020* | B.3 | 100 | 92 | 07/05/2020 | passed_qc |
| Belgium/ULG-10089/2020* | B.3 | 100 | 93 | 07/05/2020 | passed_qc |
| Belgium/ULG-10095/2020* | B.3 | 100 | 94 | 07/05/2020 | passed_qc |
| Belgium/ULG-10096/2020* | B.3 | 100 | 93 | 07/05/2020 | passed_qc |
| Belgium/ULG-10145/2020* | B.3 | 100 | 92 | 07/05/2020 | passed_qc |
| Belgium/ULG-10146/2020* | B.3 | 100 | 92 | 07/05/2020 | passed_qc |
| Belgium/ULG-10169/2020* | B.3 | 100 | 93 | 07/05/2020 | passed_qc |
| Luxembourg/LNS6264620/2020 | B.3 | 100 | 93 | 07/05/2020 | passed_qc |
| Turkey/HSGM-8992/2020* | B.4 | 100 | 100 | 07/05/2020 | passed_qc |
| Australia/VIC872/2020 | B.9 | 100 | 100 | 07/05/2020 | passed_qc |

## Notes

### Competing Interest Statement

The authors have declared no competing interest.

### Funding Statement

This work was supported by the Region Wallonne project WALGEMED(convention n 1710180).

### Author Declarations

The study was approved by the Comite d Ethique Hospitalo-Facultaire Universitaire de Liege (Reference number: CE 2020/137).

## References

1. Zhu N, Zhang D, Wang W, Li X, Yang B, Song J, et al. A Novel Coronavirus from Patients with Pneumonia in China, 2019. N Engl J Med. 2020;382: 727–733.

2. Wu F, Zhao S, Yu B, Chen Y-M, Wang W, Song Z-G, et al. A new coronavirus associated with human respiratory disease in China. Nature. 2020;579: 265–269.

3. World Health Organization. Coronavirus disease 2019 (COVID-19) Situation Report – 51. 11 Mar 2020. Available: https://www.who.int/docs/default-source/coronaviruse/situation-reports/20200311-sitrep-51-covid-19.pdf?sfvrsn=1ba62e57_10

4. World Health Organization. World Health Organization. Coronavirus disease 2019 (COVID-19) Situation Report 123. [cited 23 May 2020]. Available: https://www.who.int/docs/default-source/coronaviruse/situation-reports/20200522-covid-19-sitrep-123.pdf?sfvrsn=5ad1bc3_4

5. Drosten C, Günther S, Preiser W, van der Werf S, Brodt H-R, Becker S, et al. Identification of a novel coronavirus in patients with severe acute respiratory syndrome. N Engl J Med. 2003;348: 1967–1976.

6. Zaki AM, van Boheemen S, Bestebroer TM, Osterhaus ADME, Fouchier RAM. Isolation of a novel coronavirus from a man with pneumonia in Saudi Arabia. N Engl J Med. 2012;367: 1814–1820.

7. Chinese CDC, Specific primers and probes for detection 2019 novel coronavirus. [cited 20 Apr 2020]. Available: http://ivdc.chinacdc.cn/kyjz/202001/t20200121_211337.html

8. CDC. US CDC Coronavirus Disease 2019 (COVID-19) Real-time RT-PCR Primer and Probe. In: Centers for Disease Control and Prevention [Internet]. 10 Apr 2020 [cited 20 Apr 2020]. Available: https://www.cdc.gov/coronavirus/2019-ncov/lab/rt-pcr-panel-primer-probes.html

9. Corman VM, Landt O, Kaiser M, Molenkamp R, Meijer A, Chu DK, et al. Detection of 2019 novel coronavirus (2019-nCoV) by real-time RT-PCR. Euro Surveill. 2020;25. doi:10.2807/1560-7917.ES.2020.25.3.2000045

10. SARS-CoV-2_Sequencing. Github; Available: https://github.com/CDCgov/SARS-CoV-2_Sequencing

11. Shu Y, McCauley J. GISAID: Global initiative on sharing all influenza data - from vision to reality. Euro Surveill. 2017;22. doi:10.2807/1560-7917.ES.2017.22.13.30494

12. Hadfield J, Megill C, Bell SM, Huddleston J, Potter B, Callender C, et al. Nextstrain: real-time tracking of pathogen evolution. Bioinformatics. 2018;34: 4121–4123.

13. Dellicour S, Durkin K, Hong SL, Vanmechelen B, Martí-Carreras J, Gill MS, et al. A phylodynamic workflow to rapidly gain insights into the dispersal history and dynamics of SARS-CoV-2 lineages. bioRxiv. 2020. p. 2020.05.05.078758. doi:10.1101/2020.05.05.078758

14. Centers for Disease Control and Prevention. Implementation of mitigation strategies for communities with local covid-19 transmission. 2020. Available: https://www.cdc.gov/coronavirus/2019-ncov/downloads/community-mitigation-strategy.pdf

15. Steven Sanche, Yen Ting Lin, Chonggang Xu, Ethan Romero-Severson, Nick Hengartner, Ruian Ke. High Contagiousness and Rapid Spread of Severe Acute Respiratory Syndrome Coronavirus 2. Emerging Infectious Disease journal. 2020;26. doi:10.3201/eid2607.200282

16. Denison MR, Graham RL, Donaldson EF, Eckerle LD, Baric RS. Coronaviruses: an RNA proofreading machine regulates replication fidelity and diversity. RNA Biol. 2011;8: 270–279.

17. Artic Network. Available: https://artic.network/ncov-2019

18. Shen W, Le S, Li Y, Hu F. SeqKit: A Cross-Platform and Ultrafast Toolkit for FASTA/Q File Manipulation. PLoS One. 2016;11: e0163962.

19. Rambaut A, Holmes EC, Hill V, O’Toole Á, McCrone JT, Ruis C, et al. A dynamic nomenclature proposal for SARS-CoV-2 to assist genomic epidemiology. bioRxiv. 2020. p. 2020.04.17.046086. doi:10.1101/2020.04.17.046086

20. Vogels CBF, Brito AF, Wyllie AL, Fauver JR, Ott IM, Kalinich CC, et al. Analytical sensitivity and efficiency comparisons of SARS-COV-2 qRT-PCR assays. medRxiv. 2020; 2020.03.30.20048108.

